# Phenotypical differentiation of tremor using time series feature extraction and machine learning

**DOI:** 10.1101/2024.03.14.24303988

**Authors:** Verena Häring, Veronika Selzam, Juan Francisco Martin-Rodriguez, Petra Schwingenschuh, Gertrúd Tamás, Jan Raethjen, Steffen Paschen, Franziska Goltz, Eoin Mulroy, Anna Latorre, Pablo Mir, Rick C. Helmich, Kailash P. Bhatia, Jens Volkmann, Robert Peach, Sebastian R. Schreglmann

**Affiliations:** Department of Neurology, University Hospital Würzburg, 97080 Würzburg, Germany; Unidad de Trastornos del Movimiento, Servicio de Neurologia y Neurofisiologia Clinica, Instituto de Biomedicina de Sevilla, Hospital Universitario Virgen del Rocio / CSIC / Universidad de Sevilla, 41013 Seville, Spain; Centro de Investigación Biomédica en Red sobre Enfermedades Neurodegenerativas (CIBERNED), Instituto de Salud Carlos III, Madrid, Spain; Departamento de Psicología Experimental, Facultad de Psicología, Universidad de Sevilla, Seville, Spain; Department of Neurology, Medical University of Graz, 8036 Graz, Austria; Department of Neurology, Semmelweis University, 1083 Budapest, Hungary; Department of Neurology, Christian Albrechts University, 24118 Kiel, Germany; Department of Neurology, Donders Institute for Brain, Cognition and Behaviour, Radboud University Medical Centre, Nijmegen, the Netherlands; Department of Clinical and Movement Neurosciences, UCL Queen Square Institute of Neurology, London WC1N 3BG, UK; Departamento de Medicina, Facultad de Medicina, Universidad de Sevilla, Seville, Spain; Department of Brain Sciences, Imperial College London, London SW7 2AZ, UK

**Keywords:** tremor analysis, Essential Tremor, Parkinsońs disease, big data

## Abstract

The reliable differentiation of tremor disorders poses a significant challenge, largely depending on the subjective interpretation of subtle signs and symptoms. Given the absence of a universally accepted bio-marker, diagnostic differentiation between the most prevalent tremor disorders, Essential Tremor (ET) and tremor-dominant Parkinson’s Disease (PD), frequently proves to be a non-trivial task. To address this, we employed massive time series feature extraction, a powerful tool to examine the entirety of mathematical descriptors of oscillating biological signals without imposing bias, in combination with machine-learning (ML). We applied this approach to accelerometer recordings from tremor patients to identify the optimal recording conditions, processing, and analysis settings, to differentiate ET and PD.

We utilized hand accelerometer recordings from 370 patients (167 ET, 203 PD), clinically diagnosed at five academic centres specialising in movement disorders, comprising an exploratory (158 ET, 172 PD from London, Graz, Budapest, Kiel) and a validation dataset (9 ET, 31 PD from Nijmegen). Using 15 second recording segments from the more affected hand, we first extracted established, standardized tremor characteristics and assessed their cross-centre accuracy and validity. Second, we applied supervised ML to massive higher-order feature extraction of the same recordings to achieve optimal stratification and mechanistic exploration.

While classic tremor characteristics were unable to consistently differentiate between conditions across centres, the resulting best classifying feature combination validated successfully. In comparison to tremor-stability index (TSI), the best performing classic tremor characteristic, feature-based analysis provided better classification accuracy (81.8% vs. 70.4%), sensitivity (86.4% vs. 70.8%) and specificity (76.6% vs. 70.2%), substantially improving stratification between ET and PD tremor. Similarly, this approach allowed the differentiation of rest from posture recordings independent of tremor diagnosis, again outperforming TSI (classification accuracy 99.6% vs. 49.2%). The interpretation of identified features indicates fundamentally different dynamics in tremor generating circuits: while there is an interaction between several central oscillators in the generation of PD rest tremor, resulting in several discrete but stable signal states, signal characteristics point towards a singular pacemaker in ET.

This study highlights the limitations of current, established tremor metrics and establishes the use of feature-based machine learning as a powerful method to explore accelerometry-derived movement characteristics. More importantly, it showcases the strength of the combination of hypothesis-free, data-driven analyses and a large, multi-centre dataset. The results generated are thus resistant to device-, centre- and clinician-dependent bias and establish a generalizable differentiation method, representing a relevant step towards big data analysis in tremor disorders.

## Introduction

Tremor stands as the most frequent movement disorder ^1^, its involuntary, rhythmic, and oscillating nature predominantly affecting the hands and significantly impairing daily activities and quality of life ^2^. It is a frequent symptom of various neurological conditions, with Essential Tremor (ET) and Parkinson’s Disease (PD) being the most common, underlying aetiologies. ET, primarily a disorder of the cerebello-thalamo-cortical network, is largely believed to originate in the cerebellum ^3–5^, and is typically characterized by an isolated, bilateral postural or kinetic hand tremor ^6^. Furthermore, a considerable number of ET patients also have rest tremor^7,8^. Conversely, PD is a progressive neurodegenerative disease characterized by bradykinesia, rigidity, and a tremor that typically occurs during rest, but in many patients also re-emerges during posturing or sometimes actions ^9–12^. The pathophysiology of PD tremor is thought to involve both the basal ganglia, where tremor may be generated, and the cerebello-thalamo-cortical circuit, where the tremor is amplified, summarized in the “dimmer-switch” model ^13,14^.

Historically, clinical descriptive studies have established classic tremor phenotypes and classification systems for both disorders, thereby progressively shaping diagnostic criteria ^15–19^. However, despite these established text-book definitions, significant phenotypical and pathophysiological overlap exists ^20^, complicating clinical differentiation and leading to misdiagnoses and mistreatment in over 30% of cases ^21,22^. In response, inertial measurement units, particularly accelerometers due to their affordability, practicality, and wide availability ^23,24^, have been increasingly employed to introduce objective measures into tremor evaluation^23^, and have become standard diagnostic practice in many centres ^25–27^. ^17,18^

To date, two validated sensor-based methods to differentiate ET from PD tremor have emerged: (1) the tremor stability index (TSI) and (2) the mean harmonic power. The TSI, a metric of the relative range of frequency tolerance of a tremor, was established to compare PD *rest* tremor vs. ET *postural* tremor recordings, offering best differentiation at a TSI cut-off of 1.05 ^28^. However, absolute TSI values have been found to differ in other cohorts ^29,30^ casting doubt on its generalizability. The mean harmonic power ^31^, a metric quantifying the power at higher harmonic frequencies of the main tremor frequency, was established based on postural recordings and validated in three independent cohorts ^28,32^. However, it has been limited in its wider application due to reliance on specifically calibrated accelerometers.

Despite these advancements, established tremor metrics are unable to consistently differentiate tremor disorders ^25,33^ and a general scarcity of comparative multi-centre tremor studies further impacts the generalizability of findings. Concurrently, the use of inertial measurement devices for tremor quantification is on the rise ^34^, along with the growth in available data sets ^26^ emphasising the need for better approaches to differentiate tremors.

Machine learning (ML) describes the process of identifying patterns from data without human interference. This analytical concept is transforming medical research ^35–37^, allowing the integration and interrogation of complex data-sets by automatic pattern-recognition^29–31^. ML allows the detection of previously un-noticed, unifying, generalizable and consistent disease characteristics. In the context of accelerometric tremor measurements, numerous studies have employed ML, utilising diverse accelerometers, recording positions, algorithms, and clinical settings ^33^. However, only a minority have applied ML to the specific challenge of distinguishing between different tremor disorders ^38–42^. Moreover, issues such as limited sample sizes, monocentric study designs, and the constraints of hypothesis-driven approaches have hindered the identification of reliable, generalizable disease-specific tremor movement characteristics^33^.

Our study seeks to harness the potential of ML to uncover generalisable tremor characteristics that differentiate PD and ET patients. To capture the full spectrum of phenotypic inter-individual variability and to mitigate biases related to specific centres or devices, we combined recordings of individuals from various centres into a comprehensive, multi-centre dataset. We first explored the generalizability of established tremor characteristics. Next, by combining massive feature extraction ^43^ with ML ^44^, we identified previously unrecognized disease- and state-specific movement characteristics. Altogether, this method is advancing the accurate classification between PD and ET and represents a novel tool for hypothesis-free, unbiased time-series analysis.

## Materials and methods

### Participants and Ethics Approval

We analysed accelerometry recordings from patients with clinically established diagnoses of PD (n=203) or ET (n=167). These patients were recruited at five academic neurology institutes, each with dedicated movement disorder expertise (Graz; Budapest; London; Kiel; Nijmegen), some of which had been published previously ^31,45,46^. Clinical and demographic details were available from all centres, with the exception of the historical cohort from Kiel, and are summarized in Table 1. All patients were diagnosed by local movement disorder specialists based on at least two independent clinical consultations according to the Consensus Statement on Classification of Tremor ^17,18^. The established clinical diagnosis was used as reference throughout this work. All participants provided written informed consent according to local ethics at each participating centre prior to inclusion. The overall analysis was approved by the local research ethics committee (IRB University of Würzburg, Nr. 20210209 03) in accordance with the Declaration of Helsinki.

**Table 1).**
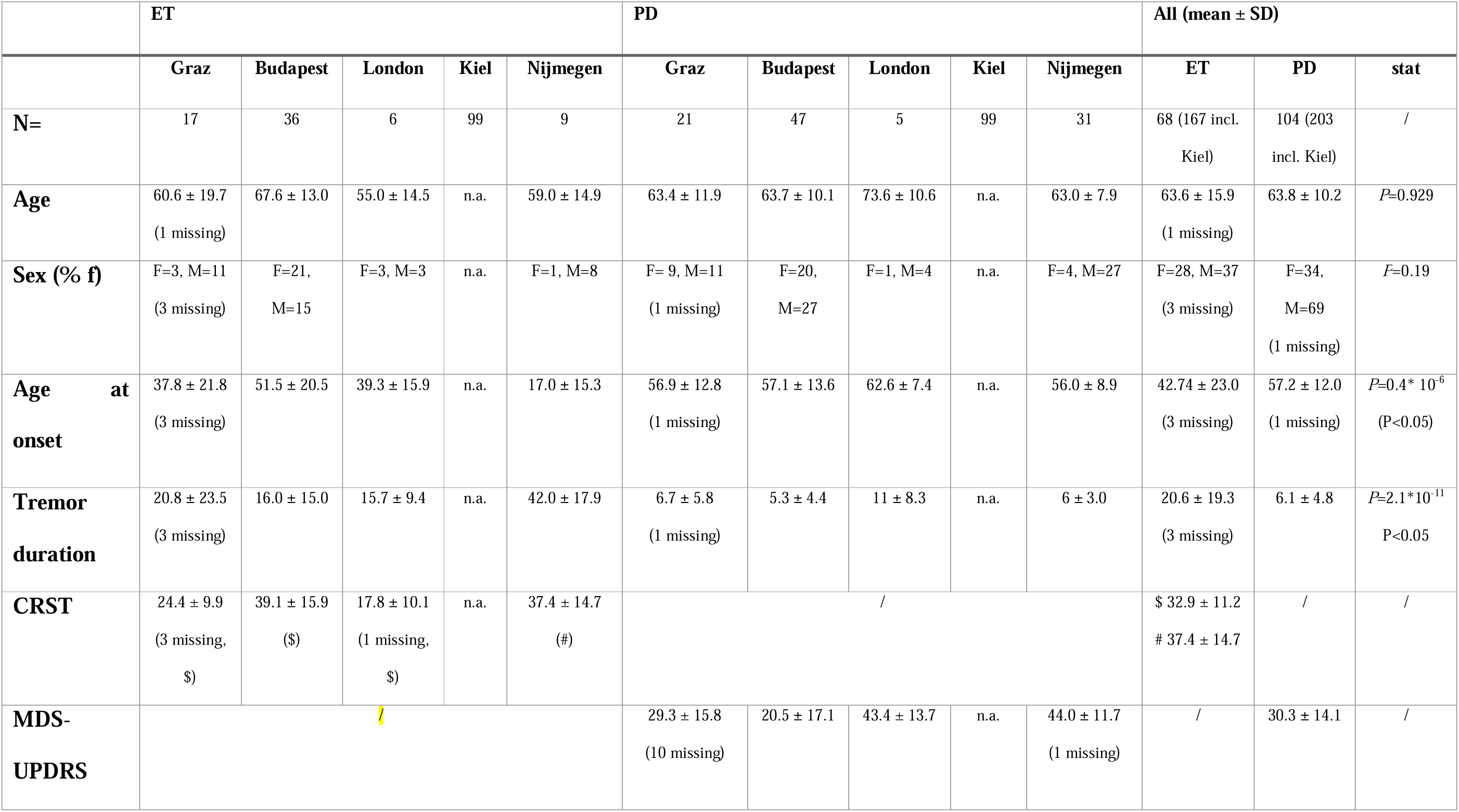
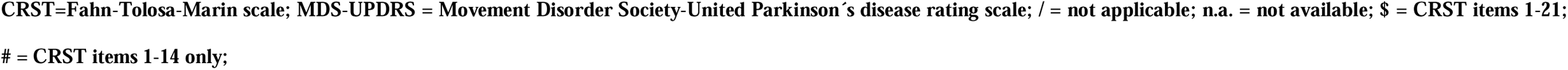
Clinical and demographic details of participants.

|  | ET |  |  |  |  | PD |  |  |  |  | All (mean ± SD) |  |  |
| --- | --- | --- | --- | --- | --- | --- | --- | --- | --- | --- | --- | --- | --- |
|  | Graz | Budapest | London | Kiel | Nijmegen | Graz | Budapest | London | Kiel | Nijmegen | ET | PD | stat |
| **N=** | 17 | 36 | 6 | 99 | 9 | 21 | 47 | 5 | 99 | 31 | 68 (167 incl. Kiel) | 104 (203 incl. Kiel) | / |
| **Age** | 60.6 ± 19.7<br>(1 missing) | 67.6 ± 13.0 | 55.0 ± 14.5 | n.a. | 59.0 ± 14.9 | 63.4 ± 11.9 | 63.7 ± 10.1 | 73.6 ± 10.6 | n.a. | 63.0 ± 7.9 | 63.6 ± 15.9<br>(1 missing) | 63.8 ± 10.2 | *P*=0.929 |
| **Sex (% f)** | F=3, M=11<br>(3 missing) | F=21,<br>M=15 | F=3, M=3 | n.a. | F=1, M=8 | F= 9, M=11<br>(1 missing) | F=20,<br>M=27 | F=1, M=4 | n.a. | F=4, M=27 | F=28, M=37<br>(3 missing) | F=34,<br>M=69<br>(1 missing) | *P*=0.19 |
| **Age at onset** | 37.8 ± 21.8<br>(3 missing) | 51.5 ± 20.5 | 39.3 ± 15.9 | n.a. | 17.0 ± 15.3 | 56.9 ± 12.8<br>(1 missing) | 57.1 ± 13.6 | 62.6 ± 7.4 | n.a. | 56.0 ± 8.9 | 42.74 ± 23.0<br>(3 missing) | 57.2 ± 12.0<br>(1 missing) | *P*=0.4* 10<sup>-6</sup><br>(*P*<0.05) |
| **Tremor duration** | 20.8 ± 23.5<br>(3 missing) | 16.0 ± 15.0 | 15.7 ± 9.4 | n.a. | 42.0 ± 17.9 | 6.7 ± 5.8<br>(1 missing) | 5.3 ± 4.4 | 11 ± 8.3 | n.a. | 6 ± 3.0 | 20.6 ± 19.3<br>(3 missing) | 6.1 ± 4.8 | *P*=2.1*10<sup>-11</sup><br>*P*<0.05 |
| **CRST** | 24.4 ± 9.9<br>(3 missing,<br>\$) | 39.1 ± 15.9<br>(\$) | 17.8 ± 10.1<br>(1 missing,<br>\$) | n.a. | 37.4 ± 14.7<br>(#) | / | | | | | \$ 32.9 ± 11.2<br># 37.4 ± 14.7 | / | / |
| **MDS-UPDRS** | / |  |  |  |  | 29.3 ± 15.8<br>(10 missing) | 20.5 ± 17.1 | 43.4 ± 13.7 | n.a. | 44.0 ± 11.7<br>(1 missing) | / | 30.3 ± 14.1 | / |

CRST=Fahn-Tolosa-Marin scale; MDS-UPDRS = Movement Disorder Society-United Parkinson's disease rating scale; / = not applicable; n.a. = not available; \$ = CRST items 1-21;

### Accelerometer examination

Across sites, accelerometer recordings were performed after an overnight withdrawal of anti-parkinsonian or anti-tremor medication. Patients were seated with their forearms fully supported to negate the effects of gravity. All patients were recorded during rest and posture bilaterally and although all recordings adhered to local practices and standard operating procedures, as described previously ^28,45,46^, slight variations existed between centres in recording positions and equipment (see Table 2). Accelerometers were attached to the dorsum of the proximal phalanx of the index (Graz, Budapest), middle finger (London), and dorsum of the hand (Kiel, Nijmegen). The recordings were tri-axial in most centres (Graz, Budapest, London, Nijmegen) and mono-axial in Kiel. Recording positions for rest were done with the hand hanging freely from the arm rest (Graz, Budapest, Kiel, Nijmegen) or the hand resting on a stable surface on its ulnar edge (London). For postural recordings, patients stretched their arms/wrists at shoulder level (Graz, Budapest, London, Nijmegen) or extended their hand while having their arms rested on the armrest (Kiel). To eliminate the influence of enhanced physiological tremor, recordings exhibiting a change in tremor frequency >1Hz upon loading with 500g/1000g attached to the wrist were excluded ^27,47^.

**Table 2).** Centre-specific accelerometry specifications.

|  | **Graz** | **Budapest** | **London** | **Kiel** | **Nijmegen** |
| --- | --- | --- | --- | --- | --- |
| **Accelerometer** | Tri-axial accelerometer<br>(Biometrics ACL300 +<br>amplifier K800, sensitivity<br>$\pm 100$ mV/g) | Tri-axial accelerometer<br>(Kinesia quantitative motor<br>assessment system; Great<br>Lakes Neurotechnologies,<br>sensitivity $\pm 4.5$ mV/g) | Tri-axial accelerometer<br>(MMA7361, Freescale<br>Semiconductor, Inc.;<br>sensitivity $\pm 800$ mV/g) | Mono-axial accelerometer<br>(Jäger-Tönnies<br>GmbH/ VIASYS Healthcare<br>Inc., sensitivity $\pm 50$ mV/g) | Tri-axial accelerometer<br>(Brain Products; sensitivity ±<br>1450 mV/g) |
| **Sampling frequency** | 1000Hz | 128Hz | 500Hz | 800Hz | 5000Hz |
| **Sensor placement** | Dorsum of middle phalanx,<br>index finger | Dorsum of middle phalanx,<br>index finger | Dorsum of proximal<br>phalanx, middle finger | Dorsum of proximal<br>phalanx, middle finger | Dorsum of the hand |
| **Rest position** | 30s hands hanging freely<br>from arm rest, supported | 15s hands hanging freely<br>from arm rest, supported | 20s hands resting on <u>ulnar</u><br><u>edge</u> on chair armrest / table | 60s hands hanging freely<br>from arm rest, supported | 60s hands hanging freely<br>from arm rest, supported |
|  | lower arm | lower arm |  | lower arm | lower arm |
| **Posture position** | 30s arms/wrists<br>outstretched at shoulder<br>level, fingers prone slightly<br>parted | 15s arms/wrists<br>outstretched at shoulder<br>level, fingers prone slightly<br>parted | 20s arms/wrists<br>outstretched at shoulder<br>level, fingers prone slightly<br>parted | 60s hands extended, prone,<br>supported lower arm | 60s arms/wrists<br>outstretched at shoulder<br>level, fingers prone slightly<br>parted |

The resulting inhomogeneity in recording positions served our primary objective: to discern disease-and posture-specific tremor characteristics from the time-series data, independent of centre-, equipment-, and position-related variables. Due to the unavailability of concurrent electromyography (EMG) recordings from every centre, EMG data could not be included in this analysis.

### Data Analysis

#### Raw accelerometer data preparation

Primary data was converted from their respective original format to a universal format for analysis in Matlab (R2021b; Mathworks, USA). All time-series raw data and its conversion in frequency and time domain (Figure 1A) were individually visualized and screened for artefacts. For tri-axial accelerometer data, the vector amplitude sum across the three available axes was calculated and used for all subsequent analyses.

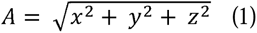

**Figure 1.**
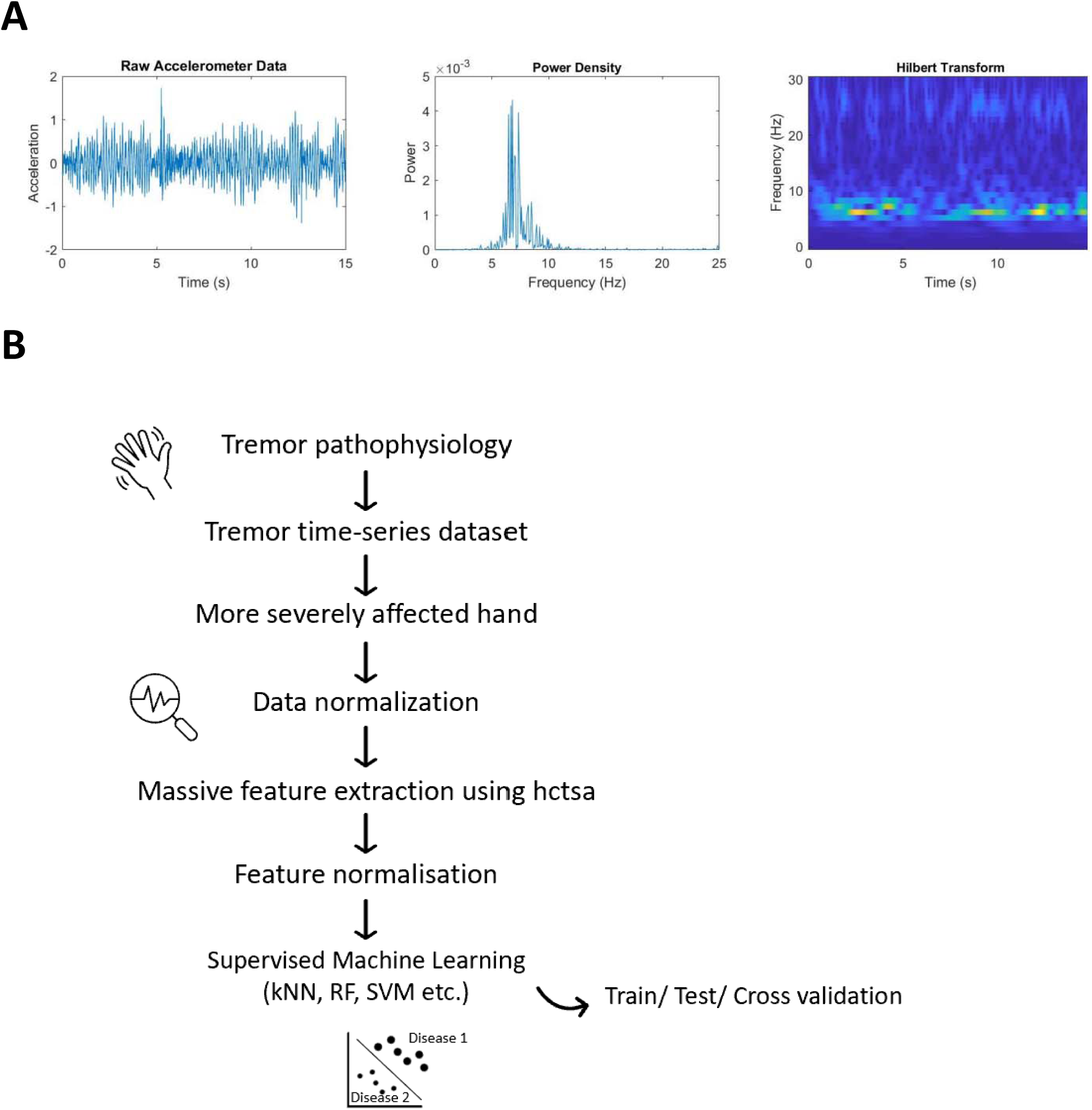
Analytical pipeline from tremulous movement and raw accelerometry signal to supervised statistical learning. (**A**) Tremor time-series data includes a wealth of information in its raw format, which can be conceptualized in frequency (power density) and time domain (Hilbert transform/wavelet). (**B**) Scheme illustrating the steps from recording tremulous hand movements via an inertial measurement unit, extraction of higher mathematical features and machine-learning to validation.

All data was normalized to a mean of 0 and standard deviation of 1, down sampled to 100Hz, standardized to a recording length of 15 seconds (at least five seconds after the onset and before the end of tremor activity to prevent artefacts) and bandpass-filtered between 2 to 30 Hz *[third-order Butterworth filter; butter and filtfilt routines in Matlab]*.

#### Detection of “more affected hand”

To further standardize analyses across data sets, and consistently use tremor recordings from the same hand for each participant throughout all analyses, we established a procedure to detect the more-affected hand, i.e. with larger tremor amplitude. Using the Area under the Curve (AUC) as a proxy for tremor power in both rest and postural position, we calculated the relative side differences in AUC for each tremor evoking position. The position with the largest relative AUC difference between sides was then used to detect the more affected hand, which was defined as the side with the larger relative AUC difference:

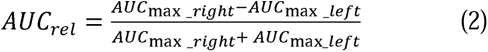

#### Standard Tremor Characteristics

Sensor-based tremor characterization so far has predominantly focused on the frequency-domain. We thus calculated the following descriptive standard tremor characteristics across recording positions, patient groups and institutes, as previously described ^28,30^: area under the curve (AUC), Tremor Stability Index (TSI), Half width power (HWP), peak frequency, full-width half maximum (FWHM), and peak power. As accelerometers and calibrations differed across centres, mean harmonic power could not be calculated/included in this work.

#### Feature-based machine-learning

To increase the scope of analytical power, we next applied an approach not limited by pre-determined characteristics, but explored the entirety of mathematical descriptors of the included n=370 time-series data, as previously established ^44^. In short, we used the highly comparative time-series analysis (hctsa) algorithm ^43,48^ to compute all possible features including autocorrelations, power spectra, wavelet decompositions, distributions, time-series models (e.g. Gaussian Processes, Hidden Markov model, autoregressive models), information-theoretic quantities (e.g. Sample Entropy, permutation entropy), non-linear measures (e.g. fractal scaling properties, nonlinear prediction errors) etc. for each time-series. This resulted in a 370 × 7873 feature matrix. After removing features with infinity, not a number (NaN) values or zero variance across the dataset this resulted in a reduced feature matrix of 370 × 7729. In order to remove the impact of variance in feature scale, the value of each feature was individually normalised to the interval [0,1] before using the feature matrix for classification.

### Model evaluation

To compare the classification accuracy of different supervised ML algorithms (TreeBagger, k-Nearest Neighbors (kNN), linear Support Vector Machine (SVM), Neural Network, and Linear Regression), data was split into train and test partitions and the balanced classification accuracy computed for each algorithm.

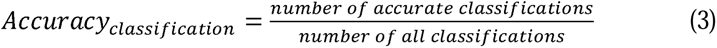

This was validated by a tenfold cross-validation, ensuring that recordings from each patient were consistently assigned to the same fold. To identify the most relevant features, we first applied a univariate approach, where features were evaluated based on their individual classification accuracies. Features exhibiting strong discriminatory power were selected for further analysis. To optimize any classification process, we next systematically tested for the influence of recording position, centre, and accelerometer axes, before testing combinations of features. After detecting relevant differences between mono- and tri-axial recordings, we optimized the classification process in all tri-axial recordings from an explorative dataset (Graz, Budapest, London) and applied this on a separate validation dataset (Nijmegen), without cross-contamination.

#### Feature Combinations

We next assessed in how far a combination of univariate features would further improve the classification accuracy. To this end, we identified features with >75% univariate classification accuracy from the exploration data set. Univariate classification accuracies were calculated across all recordings and features selected based on performance. Next, all possible combinations of two, three and four of the identified features with >75% univariate classification accuracy were assessed for their combined classification accuracy, and arranged by performance. This was done with normalized as well as non-normalized time-series data to find features dependent as well as independent of tremor amplitude.

### Comparison of recording position

In order to explore if our analysis pipeline could investigate features independent of tremor diagnosis, we determined generalizable, position-specific tremor features, that can differentiate between rest and postural recordings irrespective of tremor condition. To this end, we analysed the exploratory data set irrespective of diagnosis for differences between rest and postural recordings. Features were calculated using hctsa and the univariate classification accuracies determined. For visualization purposes the 10 best performing features were compressed into two dimensions using the Matlab implemented function *t-SNE*.

### Statistical analysis

After determining data distribution using a Kolmogorov–Smirnov Test, clinical and accelerometry data were compared using Fisher’s exact test and Wilcoxon rank sum test according to the normality distribution of data. An n-way ANOVA was applied to explore for significant differences in tremor characteristics across recording centres and diseases. All tests were two tailed, and the α level was set at p<0.05. All p values were corrected according to Bonferroni.

We used the Matlab implemented functions *[kstest2(), fishertest() and ranksum()]*as well as the statistical package JASP (JASP team 2023) for ANOVA testings.

Visualisation of multidimensional feature-space: To facilitate visualisation, principal component analysis (PCA) was performed by computing a covariance matrix for the normalised set of features. A principal component was constructed from eigenvectors and eigenvalues and displayed as a 2D scatter plot.

### Data availability

Raw data is available from individual contributing centres (according to data sharing arrangements governed by patient consent) by reasonable request to the corresponding author.

## Results

### Patient Cohorts

In this study, we analyzed data and accelerometer recordings from a total of 370 tremor patients, comprising 167 diagnosed with ET and 203 with PD. The recordings originated from different centres with detailed demographic information (Graz (17ET, 21PD) ^46^, Budapest (36ET, 47PD), London (6ET, 5PD), Nijmegen (9ET, 31PD) and a historical sample from Kiel (99ET, 99PD). None of the patients was clinically classified as ET plus, as included recordings had been collected prior to the publication of the definition of ET plus.

Among the 172 patients for whom demographic information was available, ET and PD patients were matched for age and sex. However, notable differences were observed in the age at tremor onset and mean disease duration. The onset of tremor occurred at a younger age in ET (42.72 ± 23.0 years) compared to PD patients (57.2 ± 12.0; *P*=0.4* 10^-6^) and the mean disease duration was longer in ET (20.6 ± 19.3 years) than PD patients (6.1 ± 4.8; *P*=2.1*10^-11^).

### Comparison of Standard Tremor Characteristics

We first assessed established tremor characteristics (AUC, TSI, HWP, peak frequency, FWHM, peak power) as baseline measures to benchmark differentiation accuracy. To do this, we compared tremor characteristics within and between diagnoses and across centres for rest and postural recordings (Supplementary Tables S1 & S2). Specifically, we evaluated the consistency and generalizability of these measures in differentiating ET and PD.

Our analysis revealed that rest recordings generally yielded more pronounced differences between ET and PD (Figure 2). The tremor AUC failed to consistently differentiate between the conditions, with substantial variability in measures among ET patients from different centres and contradictory trends in the two centres that showed significant differences (Figure 2A). The TSI demonstrated significant differentiation across centres overall, yet this pattern was not uniformly observed in individual centre data. Additionally, considerable variations were observed in the TSI values among ET patients from different centres (Figure 2B). The HWP achieved significant differentiation on an overall basis, but this distinction was not evident when analysing data from individual centres (Figure 2C). Peak frequency exhibited significant variability among ET patients across centres, with disease differentiation only apparent in the Kiel cohort (Figure 2D). The FWHM did not show significant differences either per disease or across centres (Figure 2E). Similar to TSI, peak power achieved differentiation between diseases across centres as a whole, yet this was only observable in two out of five centres individually (Figure 2F).

**Figure 2.**
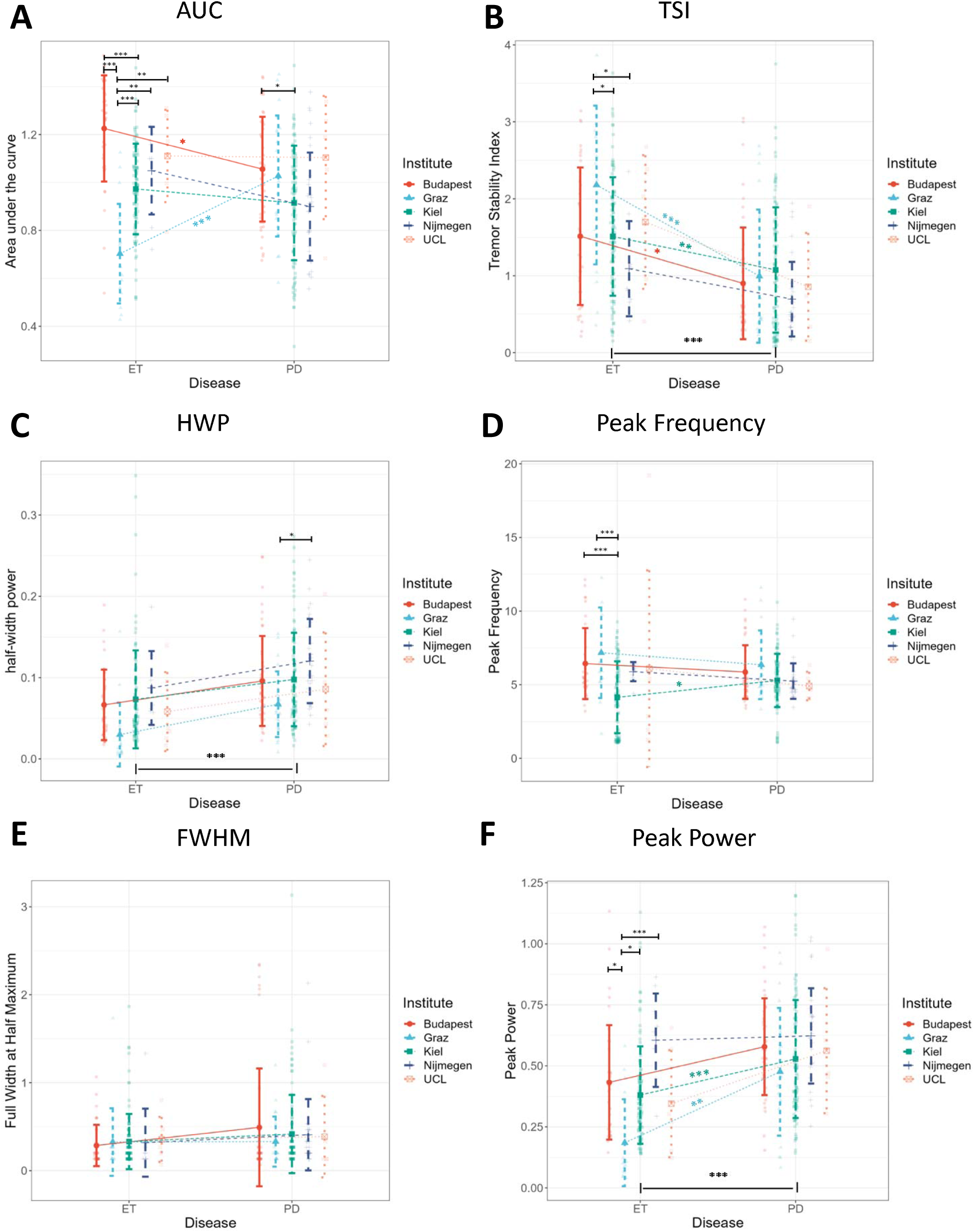
Consistency and generalizability of standard tremor characteristics in their ability to differentiate ET and PD tremor between and across centres. For the characterization of patient cohorts, established tremor characteristics including (**A**) area under the curve (AUC), (**B**) tremor stability index (TSI), (**C**) half width power (HWP), (**D**) peak frequency, (**E**) full width half maximum (FWHM) and (**F**) peak power from rest recordings were calculated and compared between centres and diseases. The comparison of half width power, peak power and tremor-stability index resulted in group differences between disorders. Notably however, while the same metrics did not detect differences between diseases in any individual centre for half width power **(C)** and for several centres for tremor stability index **(B)** and peak power **(F)**, there were substantial differences within diseases for the latter both. Hence, standard characteristics did not reliably differentiate tremor diseases in a generalizable manner.

Taken together, TSI and Peak Power provided overall differentiation between diseases, resulting in individual classification accuracies of 70.4% (TSI) and 68.9% (peak power). By combining both characteristics, classification accuracy marginally improved to 70.5%, resulting in a specificity of 80.8% and a sensitivity of 57.6% to detect a difference between ET and PD (Table 3). However, it is crucial to note that none of the standard characteristics provided a reliable and generalizable means of differentiation within this dataset.

**Table 3).**
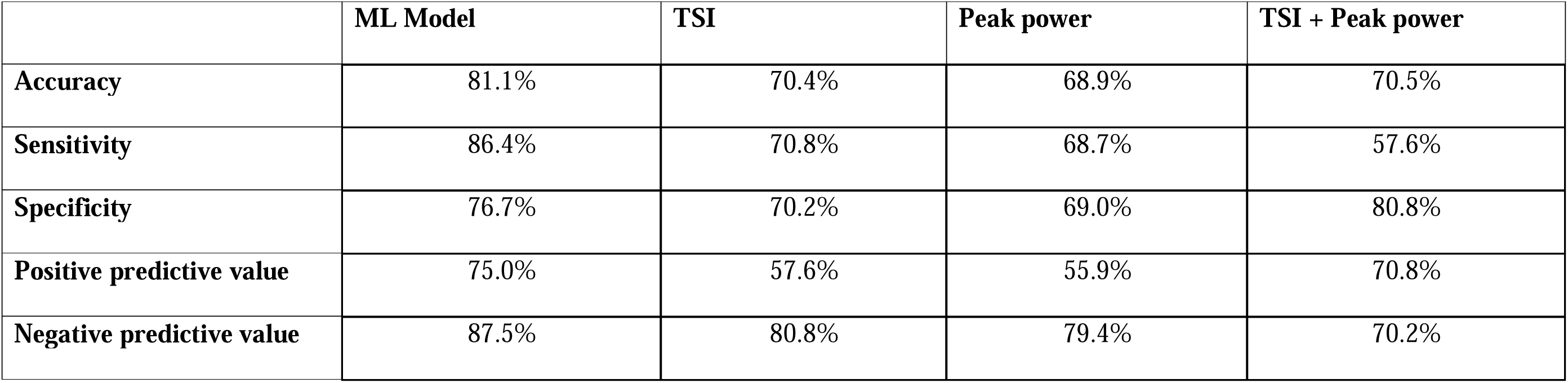
Metrics of machine-learning based classification in comparison to best-performing established tremor characteristics.

|  | **ML Model** | **TSI** | **Peak power** | **TSI + Peak power** |
| --- | --- | --- | --- | --- |
| **Accuracy** | 81.1% | 70.4% | 68.9% | 70.5% |
| **Sensitivity** | 86.4% | 70.8% | 68.7% | 57.6% |
| **Specificity** | 76.7% | 70.2% | 69.0% | 80.8% |
| **Positive predictive value** | 75.0% | 57.6% | 55.9% | 70.8% |
| **Negative predictive value** | 87.5% | 80.8% | 79.4% | 70.2% |

### Differentiation using feature-based machine-learning

#### Optimizing Recording Settings

To overcome the limitations inherent in targeted analyses, we performed a comprehensive exploration using the full spectrum of 7729 time-series features identified by hctsa using machine learning. This approach aimed to determine the optimal parameters for supervised machine learning, including the choice of algorithms, recording positions, sensor dimensionality (number of sensor axes; Figure 3A-C), and the number of top-performing features combined for classification accuracy (Figure 3A-F). This evaluation was based on one mono-centric dataset with triaxial recordings (Graz) to mitigate any centre-specific biases in recording SOPs.

**Figure 3.**
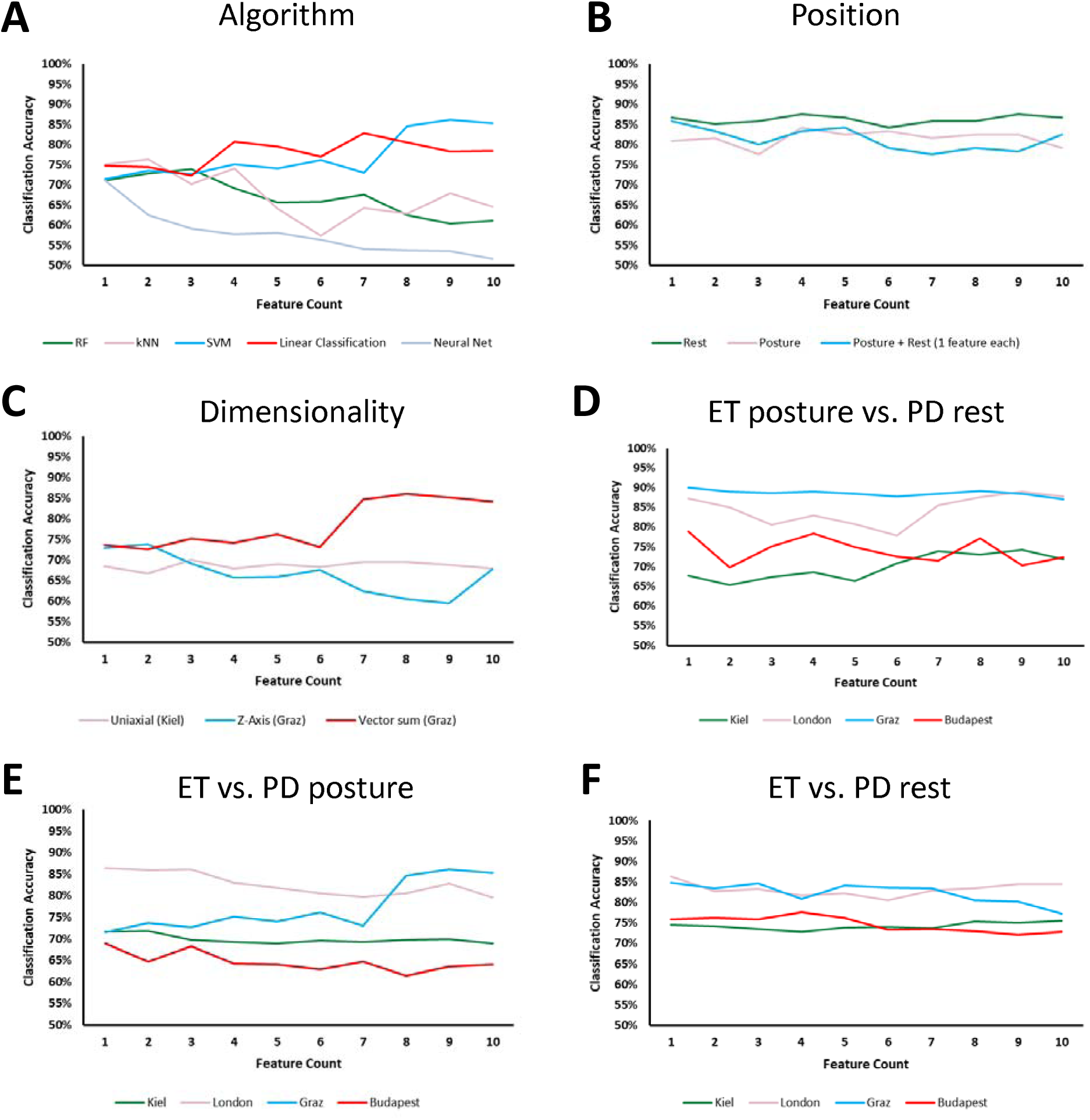
Accuracy of unbiased feature-based supervised tremor classification depends on machine-learning algorithm, recording position, sensor-dimensionality and number of features. To explore which fundamental analytical settings are best suited to differentiate ET and PD tremor, we systematically examined the effects of (**A**) machine-learning algorithm, (**B**) recording position, (**C**) sensor-dimensionality (uniaxial vs. vector amplitude sum) and number of combined best-performing features on classification accuracy. For the differentiation between PD and ET we used the comparison between (**D**) PD rest vs. ET postural, (**E**) PD vs. ET postural, as well as (**F**) PD vs. ET rest recordings. All data are based on hctsa features extracted from 15s segments of tremor accelerometer recordings (down-sampled to 100Hz) from the more severely affected hand, and support vector machine AL algorithm **(B-F**) with an ascending number of combined features on the x-axis.

We found that linear classifiers (SVM up to 86.1%, logistic regression classifier up to 82.8% classification accuracy) outperformed tree-based approaches such as Random Forest, particularly when integrating multiple features (Figure 3A). Furthermore, as above with classic tremor characteristics, we observed that recordings taken at rest consistently yielded higher classification accuracies (above 84%) compared to postural or combined recordings, irrespective of the number of features (Figure 3B).

In assessing the influence of the number of accelerometer axes on classification accuracy, we contrasted mono-axial recordings (Kiel) against both isolated single/z-axis time-series and vector amplitude sums from the same tri-axial recordings (Graz). Our findings indicated that the vector amplitude sum of triaxial accelerometer data outperformed single-axis data by approximately 13% (max. accuracy of 86.1% vs. 73.8%) and mono-axial recordings by 16% (86.1% vs. 70.0%).

Finally, we found that the combination of several best-performing features improved classification accuracy, while there was no substantial improvement beyond ten features (Supplementary Fig. 1). Taken together, we have selected SVM and vector amplitude sum generated from three axes as the ideal recording combination and tested for combinations of up to ten features in all subsequent analyses.

#### Comparison between tremor conditions

We next compared tremor recordings between ET and PD within the exploratory cohort, with a focus on how recording position influenced classification accuracy at each centre. Adopting the approach utilized in establishing the TSI ^28^, we first compared ET *postural* vs. PD *rest* recordings (Figure 3D). This approach, despite its inherent bias of contrasting different aetiology-specific postures, yielded classification accuracies as high as 90,4% for London, 89.0% for Graz, 81.1% for Budapest and 77.1% for Kiel. To address this bias, we subsequently compared ET versus PD postural recordings (Figure 3E), as well as ET versus PD rest recordings (Figure 3F). The latter comparison provided the most consistent results and was unaffected by the number of features included in the analysis.

Moreover, our findings indicated notable centre-specific variations in these analyses. For instance, recordings from Graz and London consistently demonstrated higher classification accuracies. Specifically, differentiation based on rest recordings achieved accuracies of up to 86.3% (Graz) and 84.6% (London), compared to 77.6% (Budapest) and 75.6% (Kiel). Overall, it was evident that rest tremor recordings provided more effective differentiation than postural recordings.

#### Identification of general classifiers to differentiate ET from PD tremor

We then asked whether we could identify features that reliably distinguish between ET and PD tremor signals across different centres. To do this, we analysed all available triaxial rest recordings (exploratory cohort consisting of 59 ET, 73PD) for both non-normalized as well as normalized time-series. This resulted in nine amplitude-dependent and four amplitude-independent features (Supplementary Table S3), each achieving a univariate classification accuracy of at least 75% for differentiating ET from PD tremor signals. Among these, four features were overlapping between the two categories (i.e., they appeared as top features for both the non-normalized and normalized time series) suggesting that they are independent from amplitude. Of particular note, *MF_GARCHfit* emerged as the individual feature with the highest univariate classification accuracy.

To determine the best performing combination of features across centres, we next tested all possible combinations of two, three, and four of the 13 features identified for their combined classification accuracy on all exploratory rest recordings. The paired combination of *MF_GARCHfit* and *MF_hmm_CompareNStates* emerged as the optimal choice, featuring one non-amplitude dependent and one amplitude-dependent feature. This duo yielded a combined classification accuracy of 81.8%. Importantly, the addition of more than two features did not further improve overall performance (Fig. 4A). We evaluated the performance of these two features individually and in combination across each centre - the combined use of both features consistently outperformed the use of each feature alone (Fig. 4B). Notably, *MF_hmm_CompareNStates* exhibited lower accuracy in the London dataset, but this did not significantly affect the overall combined accuracy.

**Figure 4.**
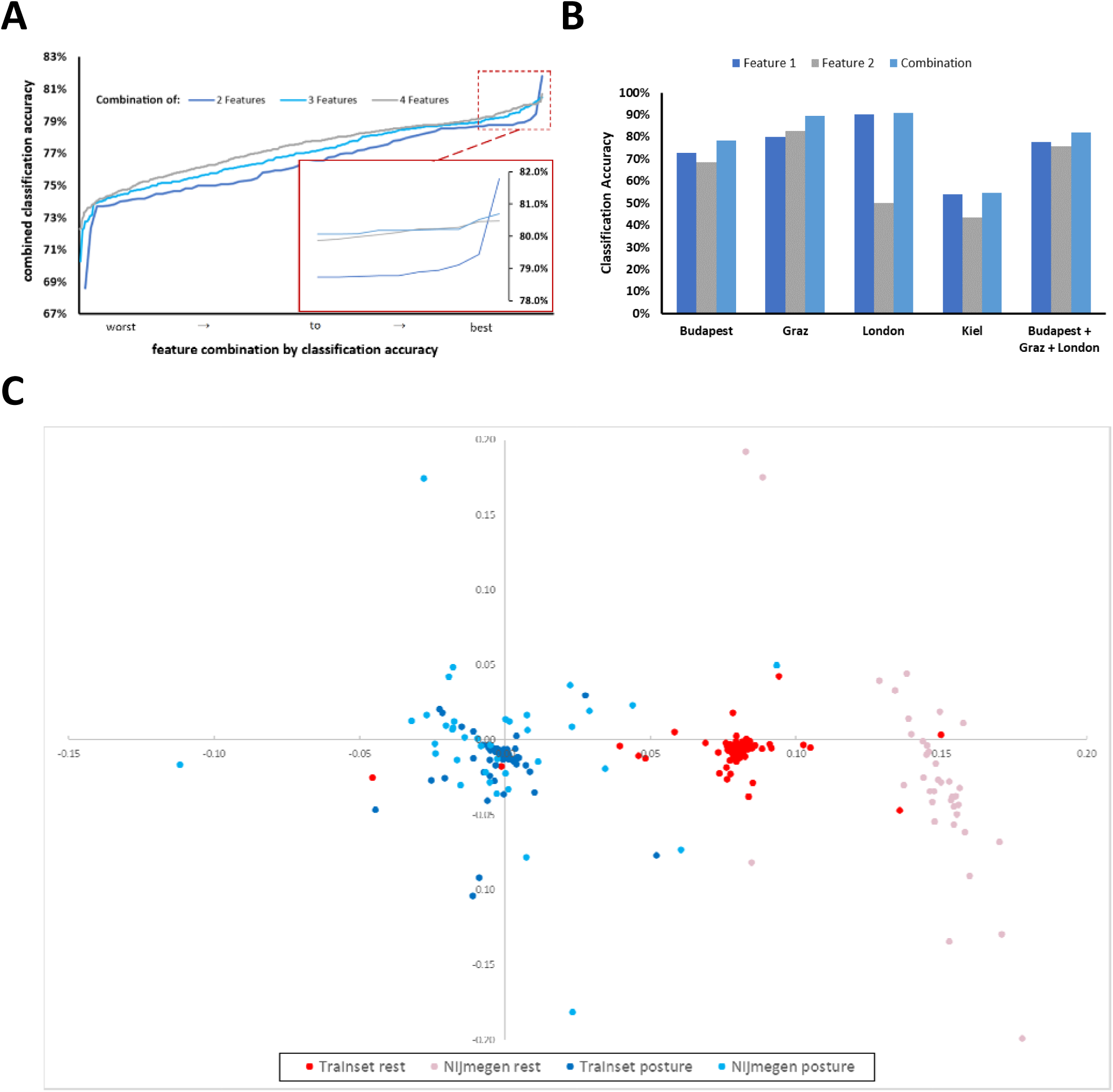
Assessment of combined, disease- and position-specific tremor features. (**A**) To identify the ideal feature combination for disease differentiation, the performance of random combinations of two, three, or four of all top-performing univariate features were assessed and arranged in ascending order (worst to best classification) by their combined classification accuracy. Notably, combining more than two features did not improve overall classification accuracy. (**B**) The two features best performing in combination (feature 1: ‘MF_GARCHfit_ar_P1_Q1.stde_normksstat’, feature 2: ‘MF_hmm_CompareNStates_06_24.maxLLtrain’) showed consistently good accuracy across tri-axial data-sets, despite feature two individually performing worse on the London data-set. Performance on mono-axial data was consistently lower. (**C**) Feature-based machine-learning also detected posture-specific tremor characteristics irrespective of clinical tremor diagnosis. The results of the ten best performing features differentiation between rest and postural recordings are combined in a multi-dimensional space and graphically displayed after dimensionality reduction (arbitrary units on both x- and y-axis). Each dot is colour-coded by dataset and represents a singular recording. The distribution of datapoints is illustrating an excellent differentiation accuracy in both the training (99.6%) and validation (97.4%) data-set, allowing reliable differentiation between rest and postural recordings irrespective of tremor diagnosis.

We also observed that these features, which were identified from tri-axial recordings across three institutes, showed more than 25% lower classification accuracy when applied to the mono-axial recordings from Kiel. This observation is consistent with our earlier findings from the standard tremor characteristic analyses, which similarly indicated diminished accuracy in mono-axial recordings.

To independently validate our model, we used the validation data set from Nijmegen (9 ET, 31 PD) and tested it with the two identified features: individually, *MF_GARCHfit* achieved 70% accuracy and *MF_hmm_CompareNStates* achieved 65% accuracy, which resulted in a combined classification accuracy of 72.5%.

### Detection of Position-Specific Tremor Features

Next, we investigated whether our feature-based ML approach could identify position-specific tremor features irrespective of tremor diagnosis. For this we compared 132 rest and 132 postural recordings from all triaxial ET and PD recordings from our exploratory cohort and calculated test metrics for established tremor characteristics as baseline. In comparison to the found poor accuracies (Supplementary Table S4), ML-based feature differentiation identified several features with excellent classification accuracy (Supplementary Table S5). Remarkably, three of these identified features achieved a perfect accuracy of 100%. Further underscoring the robustness of our approach, the combination of the ten best individually performing features yielded a classification accuracy of 99.6%, which successfully validated in the validation dataset (97.4%; Figure 4C).

### Interpretation of Disease-defining Tremor Features

To gain a more mechanistic understanding of the defining differences between ET and PD tremor characteristics, we aimed to interpret the two identified features: *MF_GARCHfit* and *MF_hmm_CompareNState* by assessing the underlying mathematics and relating it to more intuitive accelerometry data and feature distributions (Fig. 5A-D).

**Figure 5.**
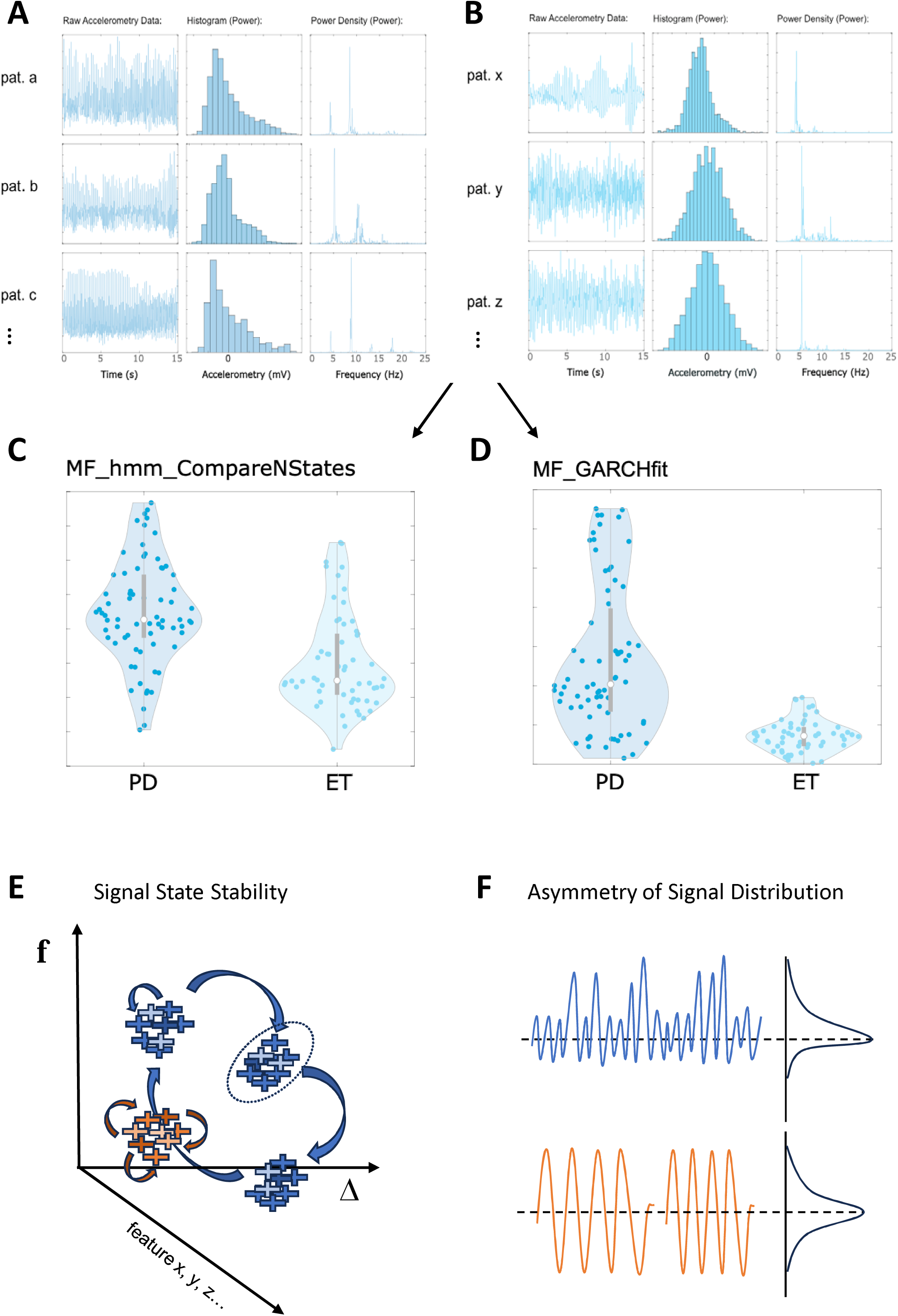
Interpretation of disease defining signal features. The two best performing features capture different aspects of the time-series signal: exemplary data plots from raw accelerometry, power histogram and power density of randomly selected **(A)** PD and **(B)** ET patient recordings graphically illustrate different signal characteristics. Violin plots capture the observed distributions and group-differences in calculated feature values between PD and ET recordings for features **(C)** ‘MF_hmm_CompareNStates’ and **(D)** ‘MF_GARCHfit’. The mechanistic translation of identified features is based on the interpretation of the underlying mathematical feature function, and graphically illustrated (**E, F**): MF_hmm_CompareNStates **(E)** assesses the position of consecutive time-series signal points (illustrated as colour-coded crosses) in a multi-dimensional feature space. Signal positions change over time, either within a ‘signal state’ (dotted circle), marking a period of relative signal stability, or between signal states, marking a change to another period of relative signal stability. ‘MF_hmm_CompareNStates’ evaluates the quality of fitting >=2 states to the data. Low quality fits of the Hidden Markov Model (HMM) suggest that the data cannot be well explained with >=2 states, suggesting that the signal stays within a singular signal state (red). Higher quality fitting of the HMM suggests that the signal naturally varies between several relatively stable signal states (blue). From the examined tremor data, ET tremor signals stay within a singular signal state, whereas PD tremor signals have a higher likelihood to change between >= 2 stable signal states. ‘MF_GARCHfit’ **(F)** examines in how far histogram-converted regular oscillations of different frequencies follow a Gaussian distribution (red) or deviate from it (blue). The latter occurs, for example, if a regular/symmetrical signal at a certain frequency is combined with asymmetrical oscillations. Higher values in this skewness metric relate to more prevalent asymmetries in the frequency distribution, as evident in PD tremor signals, whereas ET tremor signals show much lower levels of asymmetry.

We found that *MF_hmm_CompareNStates* performed well in the raw data only (not normalized time-series), suggesting that it is amplitude-dependent. This feature assesses the dynamic temporal evolution of a time-series signal in a multi-dimensional feature-space. Over time signal characteristics can remain relatively stable, i.e. within a stable ‘signal state’, or change between several ‘signal states’ (Fig. 5E). The calculation of this feature relies on a hidden Markov model as measured using log-likelihood, that is fitted to the position of signal states, with the number of identified signal states < or >= 2 states defining the feature value. The feature values of ET signals show a poor fit to >=2 states, indicating a higher probability that the signal remains within a singular signal state throughout the recording. On the other hand, in PD the data falls more clearly into several, separable but stable, generative states.

In contrast, for *MF_GARCHfit*, its presence in both normalized and raw data suggests an independence from tremor amplitude. This feature correlated with the skewness of the data, as illustrated by histogram plots (Fig. 5A, B). Specifically, this feature’s value increases with deviations from a Gaussian distribution, i.e. if the relative contributions of certain acceleration signals are asymmetrically distributed (Fig 5A). Such cases of higher signal skewness are particularly evident in PD tremor time-series (Fig. 5D). Theoretically, this asymmetry relates to signals formed by a combination of oscillations, one or more of which are characterized by a direction-specific acceleration, creating an asymmetric signal distribution (Fig. 5F). With higher feature values present in PD signals, we interpret this as a likely correlate of the more asymmetrical accelerations evident in PD rest tremor, e.g. either the pill-rolling phenomenon, or well-known sudden fluctuations in hand tremor movements.

## Stability of Features

To assess in how far our approach is influenced by recording length, we examined the stability of the identified features and their dependence on the duration of the underlying time-series. For this, we calculated the classification accuracy of the best performing combined features *MF_GARCHfit* and *MF_hmm_CompareNStates* on increasingly shortened segments of the same time-series from our exploratory cohort (59ET, 73PD; Figure 5). Our results revealed that the classification accuracy was consistently higher for feature combinations with longer time-series data. Nonetheless, even with time-series as short as two seconds, a combined accuracy of 68% was achieved, and an accuracy above 70% was achieved from a recording length of ten seconds onwards.

## Discussion

In this study, we investigated and compared established tremor characteristics, and applied massive time-series feature extraction to optimise and generalise the differentiation of ET and PD tremor accelerometer recordings. Leveraging a large, multi-centre dataset, and systematically selecting the ideal recording and analysis settings, we identified movement characteristics from time-series data that reliably replicated across centres and resulted in higher sensitivity and specificity than the best performing standard metrics TSI and peak power. Furthermore, we identified posture-specific tremor features independent of tremor diagnosis. Mechanistically, the interpretation of the identified features indicates that ET tremor accelerometer signals remains stable within an individual, singular signal state with a symmetrical signal distribution, whereas PD tremor signals can adopt >= 2 stable signal states with a more asymmetrical signal distribution.

Taken together, this work establishes multi-centre feature-based machine learning as a powerful novel method for unbiased tremor analysis. The proposed method allows reliable, valid, and generalizable differentiation of the two most prevalent tremor disorders, as well as their mechanistic exploration.

### Standard tremor characteristics provide no reliable differentiation between ET and PD tremor

This study represents the largest multi-centre tremor accelerometry investigation to date ^28^, surpassing the scale of previous research which predominantly comprised mono-centre studies ^39,41^. Our approach, combining data curation and harmonization, facilitates a nuanced interrogation of tremor disorder signals, overcoming the effects of site- or device-specific influences.

Our findings contribute significantly to understanding cross-centre variations in tremor characteristics, revealing notable differences in tremor movements among patients from different centres. While our study design does not allow us to pinpoint the exact causes of this heterogeneity – be it population, recruitment, or diagnostic bias - it arguably provides the most comprehensive overview of signal variability in ET and PD currently available. It is well-acknowledged that patient presentations at tertiary referral centres differ from those in general neurology practices, highlighting the need for future studies to encompass a broader spectrum of clinical settings.

A core observation from our data is that stratification attempts perform better with rest than with postural recordings. In fact, both TSI and peak power - the only standard metrics showing significant differences across combined data from different centres (but not consistently in each centre individually) - were found to be significant only at rest. Although not surprising – clinically ET typically does not manifest with rest tremor, except in the case of ET plus as per the latest diagnostic criteria ^18^ – this observation was consistent across classic and ML-based classification approaches. Given that ET plus was not included as a clinical diagnosis, this study cannot provide insights into the signal characteristics of this recently established clinical diagnostic category beyond published first evidence of barely discernible signal differences ^49^.

However, while both TSI and peak power demonstrated significance in combined cohort analysis, this was not uniformly observed across all individual centres, a result that could not be attributed to data from a singular centre unduly influencing the analyses. Our results hence highlight that none of the previously established tremor characteristics generalize across populations and centres – necessitating the discovery of novel metrics.

### Triaxial accelerometry is superior to mono-axial measurements

Historically, electrophysiological assessments of tremor have combined accelerometry with EMG to differentiate between the mechanical, mechanical-reflex and central component of tremor ^27^. However, over the past decade, significant advances in tremor analysis, including the development of TSI and mean harmonic peak power ^28,31^, have only utilized accelerometer data ^33,34^. While earlier studies predominantly relied on mono-axial sensors ^26,50,51^, reflecting the constraints of technology availability and cost, tri-axial accelerometers are increasingly employed ^28,33,45,46,52^ and recommended ^27^.

Our results on the effect of accelerometer dimensionality show that tri-axial accelerometer data is superior to mono-axial data in classifying tremor disorders. Mono-axial data consistently performed worse across our analyses, which cannot be attributed to sample size or centre, as exemplified in the data from Kiel and Graz. Mono-axial data hence inherently reduces the information content of the three-dimensional tremor signal, which substantially affects down-stream signal analysis. It is worth noting that while mono- and tri-axial accelerometers have been reported to be equally effective in detecting physical activity levels in adolescents and children ^53,54^, studies suggest that tri-axial sensors perform better in adults and the elderly ^55^. In line with our results, which are based on 15 second time-series data, the differentiation between types of activity over shorter time periods have been shown to rely on more fine-grained tri-axial information ^56^.

### Feature-based machine learning – an un-biased method for tremor analysis

The basic principle of ML is to “learn” patterns in complex data without human interference, offering a means to overcome limitations of classical studies, which often limit their focus to a narrow set of variables or confine their exploration to predefined, hypothesis-driven characteristics.

Our study firmly establishes feature-based ML as an unbiased, generalizable method for tremor analysis. Based on the entirety of mathematical features extracted from time-series data ^44^, this methodology incorporates the maximum available information content from high-fidelity movement recordings to achieve pattern identification and stratification. As previous efforts ^28,31^, feature-based ML is based on movement characteristics only, not taking additional clinical information into account, which is often of paramount importance in daily practice. We demonstrate that this technique nevertheless can effectively categorize tremor data by disease entity, outperforming traditional metrics. Similarly, the excellent differentiation by recording position irrespective of the clinical diagnosis confirms the analytical power of this novel approach. Our findings on ET and PD, the most prevalent tremor disorders, are derived from a multi-centre dataset incorporating phenotypic variability, limiting the vulnerability towards centre-, device- and clinician-specific bias. Results show robust performance of the identified feature-set, outperforming standard tremor characteristics.

The ongoing revolution in artificial intelligence is transforming medicine at a rapid pace and ML has been applied in several smaller tremor studies before, as reviewed by De *et al.* ^33^. The true strength of ML in this context is however only realised when paired with large, ideally multi-centre datasets, representing the physiological signal variability within a condition. By doing so in this work, we have confidently minimised *signal bias* – by taking the entirety of possible signal features into account -, *data set bias* – by combining recordings from several academic centres –, and *spectrum bias* – by aiming to sample the feature space evenly by including recordings from patients of different disease severities.

While it is anticipated that the precision of ML-based analyses will increase with larger and more clinically diverse sample sets, future international collaboration in this area is highly recommended. Nonetheless, our work demonstrates that ML-based differentiation between tremor disorders is not only feasible and superior to previous metrics but can also aid to advance our understanding of the underlying conditions.

### Deductions on Pathophysiology

The robust identification of two features describing the signal state stability (*MF_hmm_CompareNStates*) and asymmetry of signal distribution (*MF_GARCHfit*) as the defining characteristics discerning ET from PD tremor are instructive of the underlying dynamics and origin of the two tremor disorders.

Parkinsonian rest tremor is known to show spontaneous fluctuations in frequency over time and tremor amplitude is generally not affected by these subtle variations ^57,58^. This relatively broad frequency-amplitude tolerance, contrasting with a much narrower tolerance in ET, is summarized in the concept of TSI ^28^, and has been deducted from the spontaneous evolution of frequency fluctuations as well as the observation of different amplitude modulation effects despite significant frequency entrainment by brain stimulation ^57^. At the same time, tremor amplitude in PD shows a “waxing and waning” quality, and spontaneous amplitude fluctuations are preceded by increased cerebral integration several seconds before tremor onset ^59^. The detection of >= two relatively stable signal states in PD tremor time-series is strong indication of either several discrete oscillatory pacemakers, which lead the rest tremor circuitry in turn, or several dynamic activity states of the involved structures. The observation that the signal fluctuates between a discrete number of stable states points towards a dynamic interplay between involved nodes. Theoretically this could be due to the pacemaker activity undulating forth and back between structures, or alternatively effects of drifting phase alignments in the rhythmicity between core oscillatory pacemakers.

Similarly, the combination of several oscillatory signals resulting in an asymmetric signal distribution in PD tremor is incompatible with a singular pacemaker. It rather supports the current view of at least two central tremor oscillation pacemakers in PD, likely situated in the basal ganglia and the cerebello-thalamo-cortical motor loop ^60^. The currently prevailing theory on the origin of parkinsonian rest tremor, the *dimmer-switch model*, posits that the origin of tremor-related oscillations stems from the basal ganglia (globus pallidus and subthalamic nucleus), while the cerebello-thalamo-cortical loop modulates tremor amplitude^14^. Accordingly, oscillatory activity at tremor-frequency has been detected by microelectrode recordings not only in the ventral intermediate thalamic (VIM) nucleus, but also subthalamic nucleus (STN) and the internal pallidum (GPi) ^61–64^. Further, different frequency bands within the same anatomical structure (STN) have been shown to have opposing effects on PD rest tremor, representing additional, functionally discrete central oscillators ^64,65^. Our data hence provides additional evidence to support an interaction between several central oscillators in the generation of PD rest tremor, resulting in several discrete but stable signal states.

Conversely, the presence of a singular stable signal space with a symmetrical signal distribution in our data is compatible with either a singular oscillator or a much more tightly connected net of oscillators in ET. Previously, the narrower TSI has been interpreted as a result of stronger coupling between the central elements involved in tremor generation ^28,57^, and although our data cannot provide the anatomical origin of such activity, it provides a strong and direct link between tremor phenomenology and underlying pathophysiology.

### Context and Implications

Tremor disorders are diagnosed based on clinical assessment. Historically, classification systems have developed based on the clinician’s interpretation of phenotypes ^16–18,66^. This classification framework is deeply rooted in the historical routine in neurology of deducting clinico-pathological correlation from the astute observation and interpretation of subtle movement characteristics ^67^. However, the diagnostic yield of this approach has been suboptimal, particularly regarding the diagnosis of ET, which has historically been applied in a less stringent way ^68,69^.

Recent efforts to refine these classifications have led to the re-conceptualization of ET as a clinical syndrome and the introduction of ET-plus ^18^, acknowledging the diagnostic uncertainty prevalent in many tremor cases ^70^. Whilst this has been criticised for not being based on a known pathology ^71^, this new classification, which is likely yet another intermediate step in the continuing evolution of the clinical concept ET ^72^, is taking clinical soft signs into consideration. As clinical soft signs in movement disorder research are known to have low clinimetric accuracy and poor interrater reliability ^70,73–75^, however, there is a relevant clinical need for diagnostic improvement.

Despite these efforts, there remains no universally accepted biomarker to aid objective tremor classification, shrouding results from decades of research into the epidemiology, pathophysiology and genetics particularly of ET, in a cloud of ambiguity ^34,76–79^. This uncertainty must be considered when interpreting the results of studies like ours that use the current clinical diagnostic gold standard as reference.

Notwithstanding the value of clinical phenotyping, our study lays the foundation for a more objective, generalizable approach to describing and characterizing tremor. This is particularly relevant, as stratification by movement characteristics has proven to have relevant therapeutic implications, exemplified by the results from phase-locked invasive and non-invasive brain stimulation in several tremor disorders that have shown that rhythmic tremor following a sinusoidal activity is more amenable to phase-specific intervention ^44,80,81^. There is hence emerging evidence to support the notion that not the clinical diagnosis per se, but the presence of particular movement characteristics may predict the response to certain therapeutic interventions ^69,82^ – an emerging concept possibly not limited to the effect of electrical stimulation.

There is an interesting, recent debate, on whether the classic interpretation of phenotype in movement disorders should be used in a way that acknowledges the degeneracy of human movement control, i.e. the propensity of various pathophysiological entities/diseases to “break” in a limited number of phenotypes ^67^. From this, authors argued that only the reliable dynamic quantitative monitoring of movement is able to connect any specific network dysfunction with a particular phenotype. Feature-based (tremor) time-series analysis likely provides the descriptive power for any such attempts.

Several points for further improvement remain: ML approaches are known to be susceptible to incomplete sampling of the features space, e.g. by including not enough samples of common/rare variants of a disease or by failing to include populations of various genetic backgrounds. Certainly, models including larger data sets will achieve a more accurate performance and we acknowledge that the included data represent a population of predominantly European-Caucasian descent only. Outliers are another problem, as they tend to disproportionally influence ML results ^33^, and carefully curated datasets are paramount.

## Conclusion

In summary, this study establishes feature-based ML as a novel analytical tool for unbiased tremor analysis, outperforming previous electrophysiology metrics for tremor stratification, and offering a means for hypothesis-free exploration of complex time-series signals. Future studies should aim to incorporate large, diverse samples from multiple centres and various genetic backgrounds in order to further optimize the generalizability and validity of feature-based ML tremor classification.

## Supplementary material

Supplementary material is available at *Brain* online.

## Supplement

**Supplementary Fig. 1.**
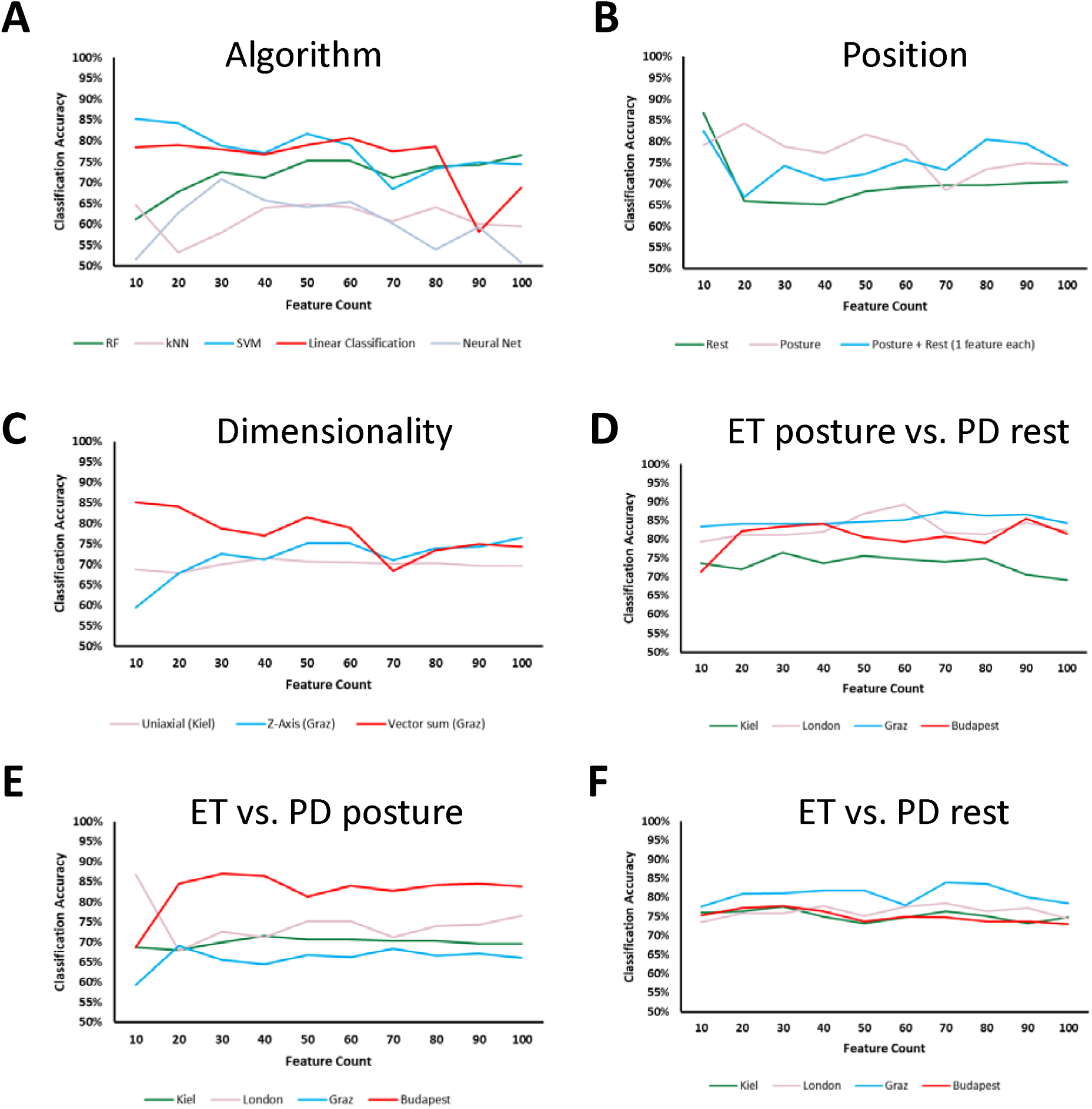
Changes of accuracy of tremor classification with ever increasing number of features. As extension of feature exploration (Fig. 3), increasing number of feature combinations were explored in order to identify the best suited feature combinations to differentiate ET and PD tremor. Extending the feature count >10 did not overall improve differentiation accuracy for the most relevant settings algorithm, position and dimensionality. All data are based on hctsa features extracted from 15s segments of tremor accelerometer recordings (down-sampled to 100Hz) from the more severely affected hand, and support vector machine AL algorithm **(B-F**) with an ascending number of combined features on the x-axis.

**Supplementary Fig. 2.**
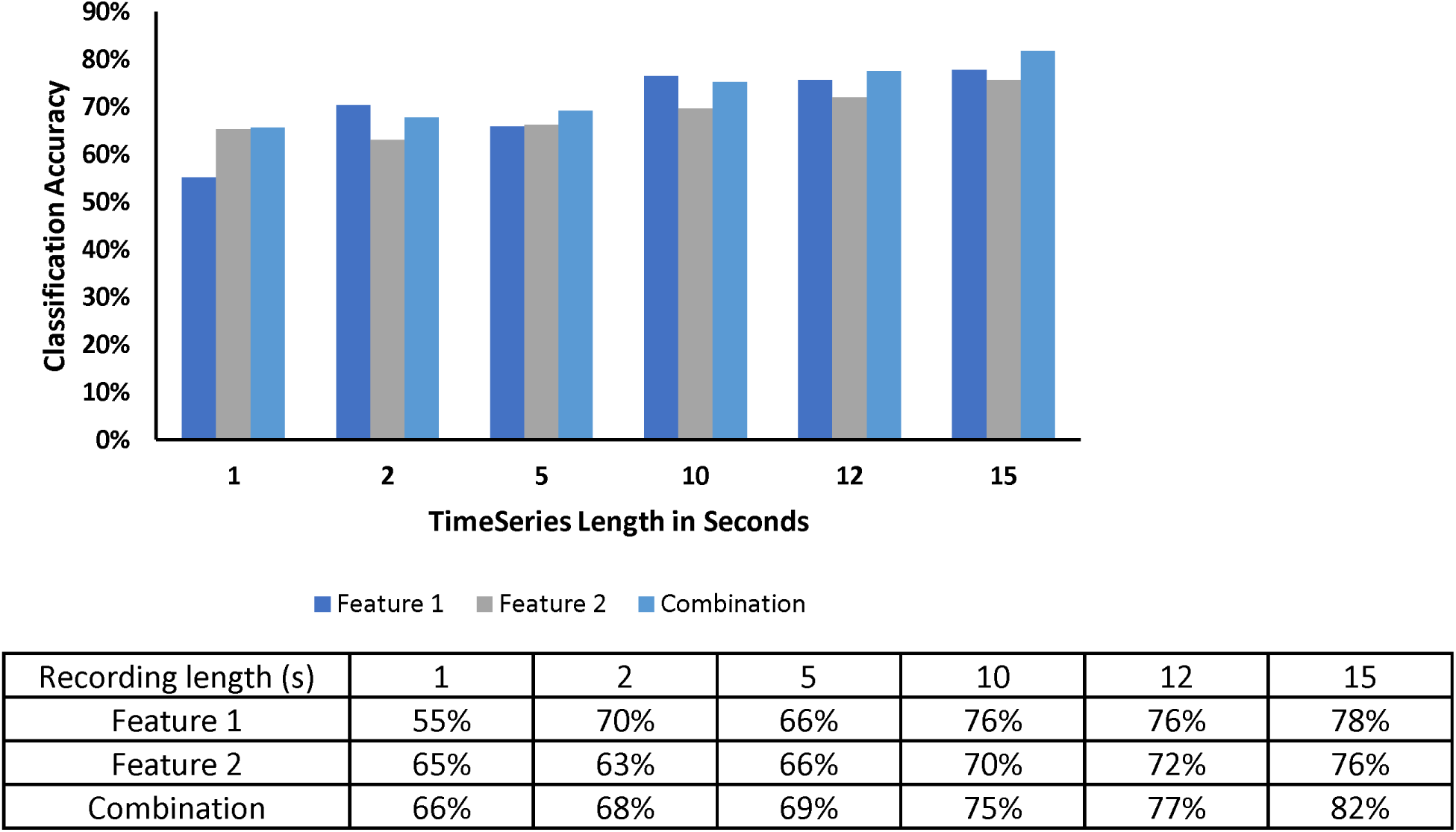
Differentiation accuracy depends on signal length. The stability and duration-dependence of features were examined using progressively longer segments of the same time-series from the exploratory cohort (59 ET, 73 PD). Differentiation accuracy gradually increased with longer time-series.

