## Supplement for "Phenotypical differentiation of tremor using time series feature extraction and machine learning"

**Table S1 Standard Tremor Characteristics – rest recordings:**

|  | ET | | | | | PD | | | | | All (mean ± SD) | |
| --- | --- | --- | --- | --- | --- | --- | --- | --- | --- | --- | --- | --- |
|  | **Graz** | **Budapest** | **London** | **Kiel** | **Nijmwegen** | **Graz** | **Budapest** | **London** | **Kiel** | **Nijmwegen** | **ET** | **PD** |
| N= | 17 | 36 | 6 | 99 | 9 | 21 | 47 | 5 | 99 | 31 | 167 | 203 |
| AUC | 0.70±0.20 | 1.23±0.22 | 1.11±0.18 | 0.97±0.19 | 1.05±0.17 | 1.02±0.25 | 1.06±0.22 | 1.10 ±0.23 | 0.91±0.24 | 0.90±0.22 | 1.01±0.24 | 0.96±0.24 |
| TSI | 2.18±1.00 | 1.51±0.88 | 1.70±0.79 | 1.51±0.77 | 1.09±0.58 | 0.99±0.84 | 0.90±0.72 | 0.86±0.63 | 1.08±0.81 | 0.70±0.48 | 1.56±0.84 | 0.96±0.76 |
| HWP | 0.03±0.04 | 0.07±0.04 | 0.06±0.04 | 0.07±0.06 | 0.09±0.04 | 0.07±0.04 | 0.10±0.05 | 0.09±0.06 | 0.10±0.06 | 0.12±0.05 | 0.07±0.06 | 0.10±0.06 |
| Peak Frequency | 7.17±2.98 | 6.44±2.37 | 6.09±6.10 | 4.14±2.43 | 5.88±0.61 | 6.36±2.27 | 5.86±1.80 | 4.91±0.87 | 5.29±1.80 | 5.25±1.18 | 5.11±2.90 | 5.52±1.80 |
| FWHM | 0.33±0.37 | 0.29±0.23 | 0.36±0.23 | 0.33±0.31 | 0.32±0.36 | 0.33±0.28 | 0.49±0.66 | 0.39±0.41 | 0.42±0.44 | 0.41±0.40 | 0.32±0.31 | 0.42±0.49 |
| Peak Power | 0.18±0.17 | 0.43±0.23 | 0.34±0.20 | 0.38±0.20 | 0.60±0.18 | 0.48±0.26 | 0.58±0.20 | 0.56±0.23 | 0.53±0.24 | 0.62±0.19 | 0.38±0.22 | 0.55±0.23 |

**Table S2 Standard Tremor Characteristics – postural recordings:**

|  | ET | | | | | PD | | | | | All (mean ± SD) | |
| --- | --- | --- | --- | --- | --- | --- | --- | --- | --- | --- | --- | --- |
|  | **Graz** | **Budapest** | **London** | **Kiel** | **Nijmwegen** | **Graz** | **Budapest** | **London** | **Kiel** | **Nijmwegen** | **ET** | **PD** |
| N= | 17 | 36 | 6 | 99 | 9 | 21 | 47 | 5 | 99 | 31 | 167 | 203 |
| AUC | 1.18±0.20 | 1.13±0.19 | 1.23±0.09 | 0.95±0.24 | 1.10±0.21 | 1.10±0.23 | 1.13±0.25 | 1.19±0.20 | 0.90±0.24 | 1.12±0.22 | 1.03±0.25 | 1.01±0.27 |
| TSI | 1.38±0.78 | 0.98±0.58 | 1.34±0.73 | 1.14±0.85 | 1.49±0.87 | 1.52±1.03 | 1.17±0.90 | 0.80±0.49 | 1.05±0.83 | 1.07±0.61 | 1.16±0.80 | 1.12±0.85 |
| HWP | 0.09±0.10 | 0.07±0.03 | 0.05±0.02 | 0.07±0.04 | 0.06±0.03 | 0.05±0.03 | 0.10±0.06 | 0.09±0.04 | 0.10±0.07 | 0.08±0.03 | 0.07±0.05 | 0.09±0.06 |
| Peak Frequency | 7.18±2.34 | 6.08±1.00 | 3.90±1.88 | 4.82±2.21 | 7.25±1.28 | 6.28±2.63 | 6.03±1.59 | 4.21±1.22 | 5.47±1.85 | 5.92±2.11 | 5.43±2.19 | 5.72±1.96 |
| FWHM | 0.55±1.10 | 0.20±0.08 | 0.27±0.09 | 0.28±0.29 | 0.17±0.06 | 0.23±0.11 | 0.47±0.66 | 0.36±0.26 | 0.40±0.59 | 0.28±0.21 | 0.28±0.43 | 0.38±0.54 |
| Peak Power | 0.41±0.20 | 0.53±0.17 | 0.39±0.11 | 0.50±0.25 | 0.58±0.27 | 0.35±0.18 | 0.50±0.16 | 0.56±0.25 | 0.54±0.23 | 0.50±0.17 | 0.50±0.23 | 0.51±0.21 |

**Table S3 List of individually best-performing time-series features to differentiate ET from PD tremor recordings; feature 1-9 are amplitude-dependent, feature 10-13 are amplitude-independent;**

| **Feature-number** | **Feature:** | **Univariate Classification Accuracy:** |
| --- | --- | --- |
| **Amplitude-dependent:** | | |
| 1046 | 'NL_DVV_3_100_2_50_10_default.trend' | 75.13% |
| 1169 | CO_AddNoise_1_quantiles_10.firstUnder50 | 75.13% |
| 1208 | 'CO_AddNoise_1_even_10.firstUnder75' | 75.13% |
| 2858 | EN_mse_1-10_2_015_diff1.meanch | 75.59% |
| 6581 | WL_dwtcoeff_db3_5.noisestd_l1 | 75.13% |
| 6713 | NL_MS_nlpe_2_mi.normp | 75.03% |
| 7584 | MF_hmm_07_3.LLtrainpersample | 75.79% |
| 7601 | MF_hmm_CompareNStates_06_24.meanLLtrain | 77.13% |
| 7602 | MF_hmm_CompareNStates_06_24.maxLLtrain | 75.69% |
| **Amplitude- independent:** | | |
| 7441 | MF_GARCHfit_ar_P1_Q1.stde_normksstat' | 77.79% |
| 7442 | MF_GARCHfit_ar_P1_Q1.stde_normp | 78.67% |
| 7498 | MF_GARCHfit_ar_P1_Q2.stde_normksstat | 77.23% |
| 7499 | MF_GARCHfit_ar_P1_Q2.stde_normp | 77.23% |

**Table S4 Metrics of machine-learning based classification for comparing rest from postural tremor recordings in comparison to established tremor characteristics.**

|  | **ML Model** | **AUC** | **TSI** | **HWP** | **Peak frequency** | **FWHM** | **Peak power** |
| --- | --- | --- | --- | --- | --- | --- | --- |
| **Accuracy** | 99.6% | 39.0% | 49.2% | 43.2% | 48.5% | 45.5% | 47.0% |
| **Sensitivity** | 99.2% | 48.5% | 59.1% | 43.2% | 64.1% | 43.9% | 38.6% |
| **Specificity** | 100% | 29.5% | 39.4% | 43.2% | 35.6% | 47.0% | 55.3% |
| **Positive predictive value** | 100% | 40.8% | 49.4% | 43.2% | 48.8% | 45.3% | 46.4% |
| **Negative predictive value** | 99.2% | 36.4% | 49.1% | 43.2% | 48.9% | 45.6% | 47.4% |

**Table S5 Top 10 individually best-performing tremor features to differentiate rest- from postural tremor recordings irrespective of clinical diagnoses (ET or PD); features are amplitude dependent;**

| **Feature Number:** | **Feature:** | **Univariate Classification Accuracy** |
| --- | --- | --- |
| 1036 | HT_DistributionTest_chi2logn5 | 100% |
| 1218 | CO_AddNoise_ac_quantiles_10.linfit_mse | 100% |
| 1256 | CO_AddNoise_ac_even_10.linfit_mse | 100% |
| 5270 | NL_TSTL_dimensions_50_ac_fnnmar.scr_bc_m2_meansqres | 99.6% |
| 7561 | SY_VarRatioTest_4_1.ratio | 99.2% |
| 7567 | SY_VarRatioTest_24682468_00001111.IIDperiodmaxpValue | 99.2% |
| 5975 | PP_Compare_spline64.olbt_s2 | 59.2% |
| 6254 | PP_Compare_rav10.olbt_s2 | 58.9% |
| 5826 | PP_Compare_sin1.statav10 | 56.6% |
| 6012 | PP_Compare_diff2.swss10_1 | 55.4% |
